# The Common Interests of Health Protection and the Economy: Evidence from Scenario Calculations of COVID-19 Containment Policies

**DOI:** 10.1101/2020.08.14.20175224

**Authors:** Florian Dorn, Sahamoddin Khailaie, Marc Stoeckli, Sebastian C. Binder, Berit Lange, Stefan Lautenbacher, Andreas Peichl, Patrizio Vanella, Timo Wollmershäuser, Clemens Fuest, Michael Meyer-Hermann

**Author notes:** First authors in alphabetic order (Florian Dorn, Sahamoddin Khailaie, Marc Stoeckli).

## Abstract

Several countries use shutdown strategies to contain the spread of the COVID-19 epidemic, at the expense of massive economic costs. While this suggests a conflict between health protection and economic objectives, we examine whether the economically optimal exit strategy can be reconciled with the containment of the epidemic. We use a novel combination of epidemiological and economic simulations for scenario calculations based on empirical evidence from Germany. Our findings suggest that a prudent opening is economically optimal, whereas costs are higher for a more extensive opening process. This rejects the view that there is a conflict with health protection. Instead, it is in the common interest of public health and the economy to relax non-pharmaceutical interventions in a manner that keeps the epidemic under control.

---

The SARS-CoV-2 virus pandemic confronts the world with a rapid spread of infections and deaths associated with COVID-19. Several governments use(d) non-pharmaceutical interventions (NPIs) to slow down and contain the epidemic. These strategies include social distancing measures, prohibition of public events, curfews, school closures, and restrictions of business activity. Evidence suggests that these measures indeed reduce the number of infections^1,2^. At the same time, the shutdown measures give rise to substantial economic costs^3,4^.

In the public debate on the further course of action, interests of public health and the economy are often presented as being in conflict^5,6^. Although this trade-off view may seem intuitive, the underlying situation is more complex. The simple trade-off view neglects that an unregulated spread of the virus with larger disease burden would also have adverse effects on the economy^7,8^. Evidence on medium- and long-run economic consequences of past pandemics supports this view^9–11^. New infection waves, e.g. due to accelerated loosening of restrictions, would reduce trust of consumers and investors, and companies would have to shut down their business activities again – regardless of government regulations – resulting in considerable further costs. Conversely, stricter regulation may also give rise to indirect disease burden in other areas. The aim is to make the fight against the pandemic sustainable^12^ and to reconcile public health and economic objectives.

An increasing number of studies examines NPI strategies with respect to epidemic and economic consequences, oftentimes concluding that partial shutdowns and immediate health policy interventions may be the most favorable strategy^8,11,13^. A central question in the public debate, however, is about the optimal exit strategy out of the shutdown. Policy-makers must weigh the extent of the restrictions against economic consequences. A conflict between health protection and economic interests could arise if the strategy with lower economic costs would lead to significantly higher COVID-19 deaths. Such a conflict would be particularly relevant if the reduction of economic costs required a rapid opening process. Using Germany as a case study, we examine the economically optimal exit strategy which can be reconciled with further containment of the epidemic.

Germany is currently discussed as a best practice example from an international perspective. Like many other countries, Germany introduced restrictive shutdown measures in several stages in March 2020 to contain the spread of the virus. In addition, many firms and citizens adapted their behavior and largely complied with social distancing policies. The sum of these measures and behavioral changes influenced the effective reproduction number in Germany. *R*_t_ fell well below one in April^14^, and the number of daily new reported cases decreased noticeably during the shutdown (see Fig. 1). On April 20^th^, a gradual loosening of the restrictions was announced. With a lag of two weeks – due to the time it takes for symptoms onset and case reporting – *R*_t_ increased again at the beginning of May (see Fig. 1).

**Figure 1:**
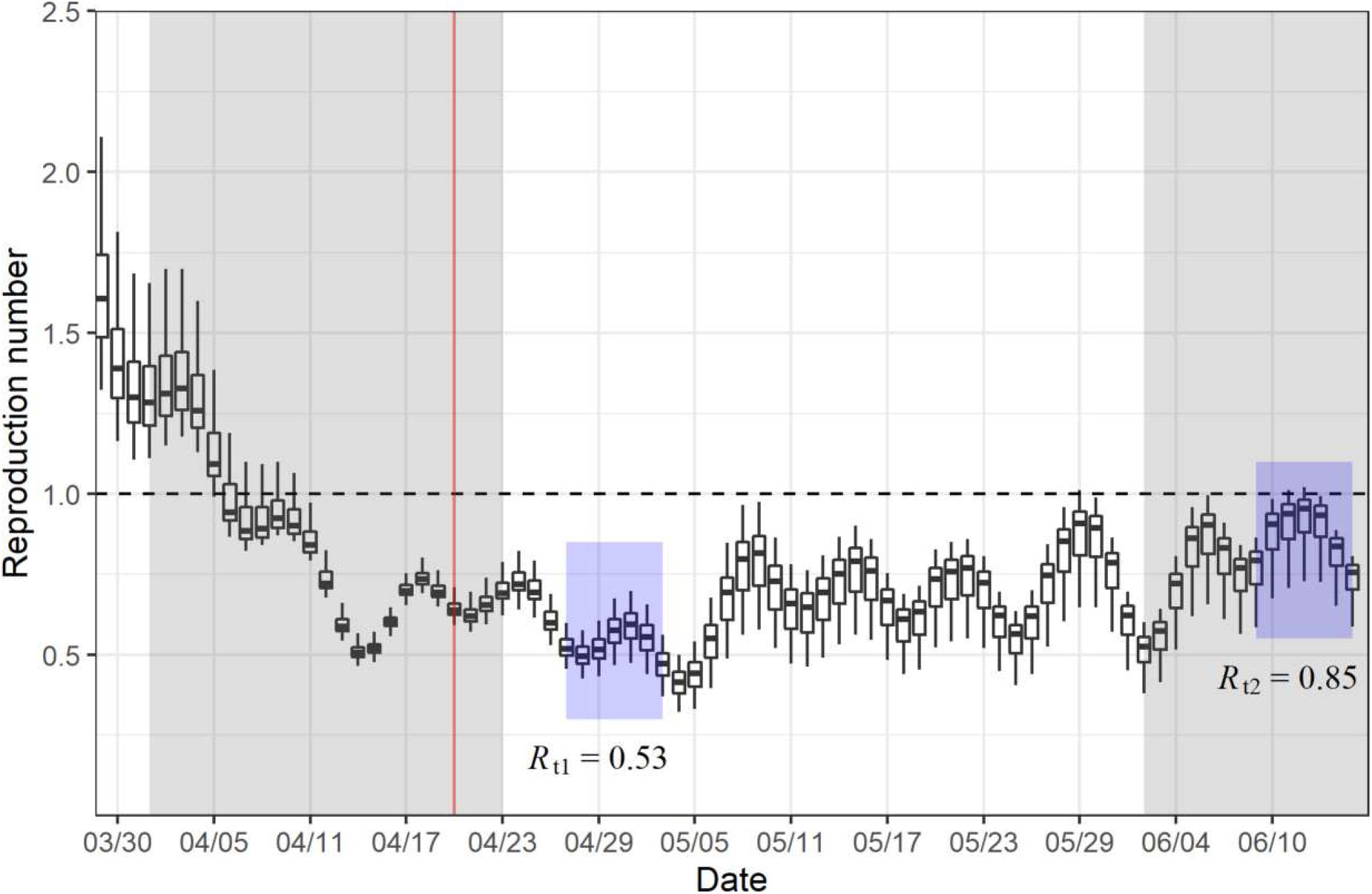
Reproduction number in Germany over time. For each date, the boxplots illustrate the distribution of the *R*_t_ estimates (median, 25 and 75 percentile). The error bars denote 1.5 times the interquartile range. The shaded grey areas indicate the survey periods for the ifo Business Survey in April and June. The shaded blue areas indicate the time window that was used to calculate the reference *R*_t_ values in the status quo before and after the impact of gradual lifting of NPIs. The vertical red line indicates the beginning of the gradual lifting of NPIs in Germany on April 20^th^.

## Combining epidemiology and economy to estimate impact of opening NPIs

The status quo of our scenario calculations represents the situation of *R*_t_ and economic activity in the initial shutdown phase until the gradual opening process started on April 20^th^. Starting from the status quo, we simulate various scenarios for further loosening or tightening the shutdown measures. We model the death toll and economic activity as a function of *R*_t_, using an empirical relationship between *R*_t_ and economic activity at the industry level as well as the time until the economy fully recovers. In the model, different shutdown strategies are associated with different *R*_t_ values; more relaxed (restrictive) restrictions yield larger (smaller) *R*_t_ values, implying a longer (shorter) period until the containment of the epidemic is completed. A longer period is associated with larger death tolls, but due to more relaxed restrictions also with higher economic activity in the short run. However, larger *R*_t_ values imply that the time until the level of new infections allows a full opening of the economy is extended. That way, it is *a priori* not clear which strategy is economically optimal in the long run.

Our scenario calculations are based on a novel and unique combination of epidemiological and economic simulation models. In a first step, we employ a mathematical-epidemiological model with Susceptible-Exposed-Carrier-Infected-Recovered (SECIR) components to estimate the development of *R*_t_ in Germany^14^ (see Fig. 1, and supplement). The estimates are based on a dynamic adaptation of the model parameters to the incidence reporting database of the Robert Koch Institute (RKI), the German government’s central scientific institution for monitoring the situation on SARS-CoV-2. We specify *R*_t_1 = 0.53 as reference value that refers to the estimated reproduction number in Germany just before the partial lifting of the NPIs on April 20^th^ (Fig. 1, left blue area). Similarly, we specify *R*_t_2 = 0.85 as reference value that refers to the reproduction number after the partial lifting, thus capturing the effect of lifting the NPIs on *R*_t_ (Fig. 1, right blue area).

## COVID-19-associated death tolls are stable at intermediate reproduction numbers

Germany counts 4,404 registered COVID-19 deaths as of April 20^th^, 2020. The calibrated simulation model projects the number of expected additional COVID-19 deaths until July 31^st^, 2021. The death toll rises with increasing reproduction numbers, although the differences are relatively small up to *R*_t_ = 0.75 and stay within a range of 10% additional deaths relative to the reference scenario with *R*_t_ = 0.53 (Fig. 2A). In contrast, the death toll rises sharply from *R*_t_ = 0.9 onward. Expected additional deaths increase to around 300% compared to the reference scenario at *R*_t_ = 1.0.

**Figure 2:**
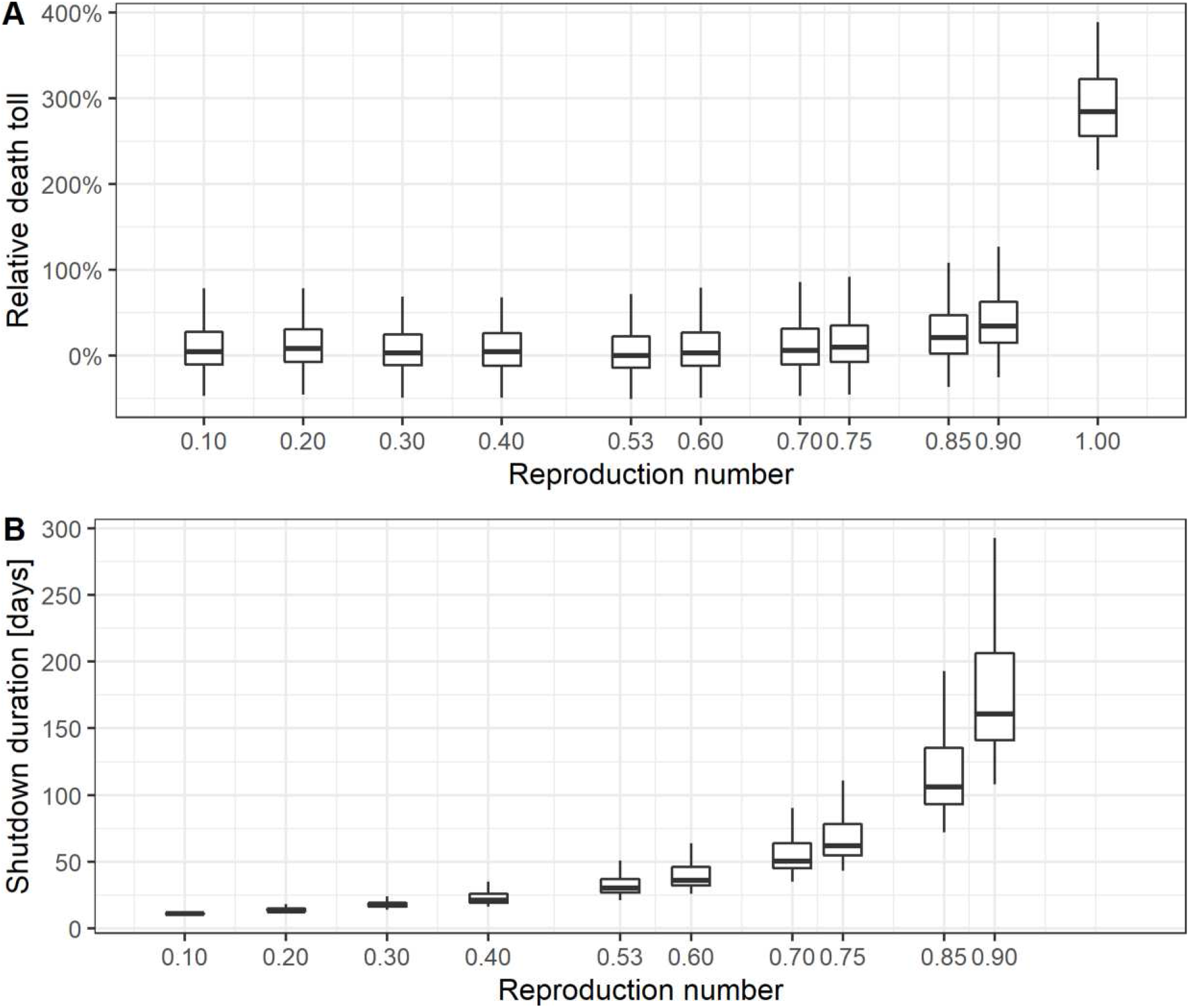
**(A)** Estimation of the relative death toll accumulated between April 20^th^, 2020 and July 31^st^, 2021 with the epidemic model, in percentage difference to the median value in the reference scenario (*R*_t_ = 0.53). The reproduction number on the abscissa was fixed in the simulation from April 20^th^, 2020 until reaching 300 daily new cases per day and then set to one. **(B)** Estimation of the shutdown duration needed to reach 300 new reported cases per day for each fixed reproduction number, starting on April 20^th^. The boxplots illustrate the distribution of the estimates (median, 25 and 75 percentile). The error bars denote 1.5 times the interquartile range.

## A larger reproduction number delays control of the epidemic

A key assumption in our analysis is that reducing the number of new infections to 300 reported cases per day would allow to control the epidemic through tracing by the approximately 400 public health offices in Germany. The remaining restrictions limiting economic activities could be lifted thereafter. In a prospective study, we assume different values of *R*_t_ and keep the value constant until the threshold of 300 daily new infections per day is reached, determining the duration of the shutdown. The fewer restrictions are imposed, the longer the restrictions need to be kept in order to reach the threshold (Figure 2B). Importantly, the reproduction number impacts nonlinearly onto the time-period required to reach the threshold. In addition, we consider a scenario where *R*_t_ is kept at one until a vaccine is generally available. While this strategy would not aim at further reducing the number of new infections but keeping it at a constant level, it allows for more extensive loosening of restrictions. In the baseline scenario, we assume that the vaccine becomes available at a large-scale on July 31^st^, 2021.

## Recovery of economy depends on the speed of opening NPIs

In a second step, we integrate the results from the SECIR model into the economic model and simulate the economic costs for different policy scenarios. The costs of a scenario are given as the aggregated loss of economic activity during the shutdown and recovery period. Taking into account that the shutdown has a different impact across industries, we explicitly model activity at the level of industry sectors. Overall economic activity is then given by the weighted sum of the industry-specific activity levels, where the weights refer to the industry’s share in total output.

Figure 3A illustrates the process in the economic model for the scenario where the policymakers increase *R*_t_ from 0.53 to 0.85 on April 20^th^. This refers to the change in the reproduction number and economic activity that was empirically observed over that period of time in Germany. The model assumes that the economy starts from a pre-shutdown activity level and experiences a decline due to the shutdown imposed in March 2020. Activity levels of different industries in response to the shutdown are determined by the status quo before the exit process started on April 20^th^. Prior to reaching the required 300 new cases per day, our model assumes that policy-makers can decide on the further course of severity of restrictions in a period of partial opening. Loosening restrictions would ceteris paribus increase economic activity in the partial opening phase (see Fig. 3A). But it would also give rise to higher *R*_t_ values, thus increasing the duration of this phase (see Fig. 2B). Conversely, tightening restrictions would lead to a reduction in economic activity, but reduces *R*_t_ and the time needed until the number of new infections would allow a full opening. The economy slowly recovers once all shutdown measures can be fully lifted without jeopardising the containment of the epidemic because either a sufficient low number of new infections is reached or a vaccine is available. At the end of the recovery phase, the economy returns to its pre-shutdown activity level.

**Figure 3:**
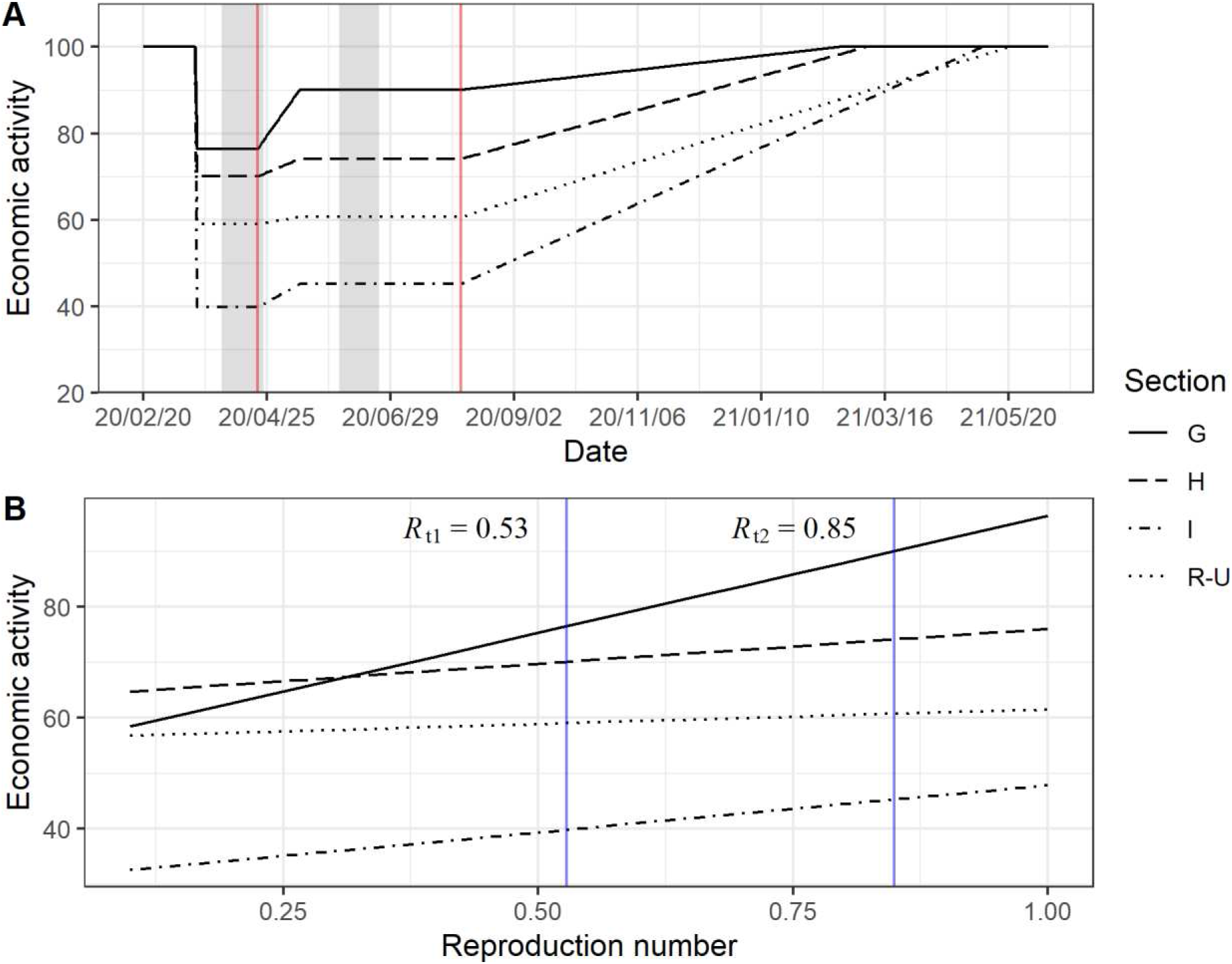
**(A)** The process of economic activity for the scenario where the policymakers increase *R*_t_ from 0.53 to 0.85. Starting from the pre-shutdown level (normalized to 100), the economy experiences a decline in activity during the shutdown. On April 20^th^, the policy-makers initiated a gradual lifting of NPIs (indicated with the first vertical red line). After the 300 daily new cases have been reached, the measures are lifted and the economy enters the recovery phase (indicated with the second vertical red line). The beginning of the recovery phase depends on the *R*_t_ value and the associated time period in Fig. 2B). Depicted are the activity levels for the economic sections *G* (wholesale and retail trade), *H* (transportation and storage), *I* (accommodation and food service activities), and *R* to *U* (entertainment and other service activities). The shaded grey areas indicate the survey periods for the ifo Business Survey in April and June. A more in-depth description of the model can be found in the supplement (see Fig. S4). **(B)** The linear relationships between changes in industry-specific economic activity and changes in the reproduction number. The vertical blue lines indicate the *R*_t_ values 0.53 and 0.85 that are used to estimate the slope.

The estimates of economic activity are based on the ifo Business Survey, a monthly survey that includes roughly 9,000 responses from German firms in manufacturing, the service sector, trade, and construction^15^. The survey statistics provide leading indicators for the German economy and are available more timely than official data. We use the companies’ assessment of their current business situation as it is highly correlated with the gross value added and several official economic activity measures^16^. We relate firm responses during the survey periods to different shutdown and partial opening periods as well as the corresponding *R*_t_ values from the SECIR model (see Fig. 1). Specifically, we assume a linear relationship between *R*_t_ and changes in economic activity for each industry using the April and June surveys and the corresponding reference reproduction number values, *R*_t1_ = 0.53 and *R*_t2_ = 0.85. The April survey captures the activity levels in each industry during the shutdown before partially lifting restrictions, whereas the June survey captures the activity levels after the lifting process.

## The depression of economic activity is sector-specific

Figure 3B shows the industry-specific linear relationship between the reproduction number and economic activity, and illustrates that elasticities in economic activity differ across industrial sections. For example, lifting shutdown restrictions gives rise to a larger increase of economic activity in retail (*G*) compared to transportation industries (*H*).

Not all industries in Germany were directly affected by legal shutdown measures. The manufacturing industry or electricity firms, for example, were not included in any state order to shut down their business activities in Germany. However, these industries also experience large slumps in activity because of declined demand from domestic and abroad, disrupted supply chains, or absences from work due to illness and quarantine. We thus distinguish between exogenous and endogenous industries in our simulation model (see Table S2 in the supplement). The former refers to the industries that are exogenously shut down by the government and are treated as illustrated in Figure 3B. These include, amongst others, firms in retail trade, accommodation and food services, transportation, entertainment and recreation, and several other service activities. Changes in activity levels in endogenous industries such as the manufacturing sector, however, are only affected by changes in the economic output of exogenous industries. We exploit inter-industry linkages and use input-output tables for the German economy to specify to what extent the activity level in one industry is affected by changes in other industries^17^.

We introduced a special question in the ifo Business Survey. Respondents were asked about the expected duration until their business situation would return to normal once all shutdown measures are lifted. For the reference scenario (*R*_t_ = 0.53), we take the mean of these expectations for each industry (see Table S2 in the supplement). We assume that it takes the firms two months less to fully recover in the scenario with *R*_t_ = 0.85 compared to the reference scenario. That way we construct a data-based linear relationship between *R*_t_ and the time to recover to the pre-shutdown level for different industries (see recovery phase in Fig. 3A).

## The long-term economic costs are minimal at intermediate reproduction number

The total economic costs of the scenarios result from the aggregated loss of economic activity among all industries over the years 2020-2022. Figure 4A shows the development of aggregated activity in our policy scenarios as deviations from the pre-shutdown activity level (normalized to 100). The scenario with *R*_t_ = 0.1, i.e. where the policy-makers further intensify the shutdown, would further decrease economic activity. On the other hand, lifting the restrictions such that the reproduction number increases to one gives rise to larger activity thereafter. However, some restrictions have to be kept in place for such a long time that the pre-shutdown activity level is not reached before 2022. Figure 4B shows the relative costs for all scenarios compared to the reference scenario (*R*_t_ = 0.53). The relative costs are given as the percentage differences in total loss of economic activity. The results show that both a strategy with extending high levels of restrictions (*R*_t_ < 0.53) and a strategy with too aggressive loosening of measures (*R*_t_ > 0.9) would lead to higher relative economic costs. Compared to the strategy of keeping restrictions of the initial shutdown period (*R*_t_ = 0.53), costs decrease in a strategy of a slight loosening of restrictions. The optimal reproduction number is around 0.8.

**Figure 4:**
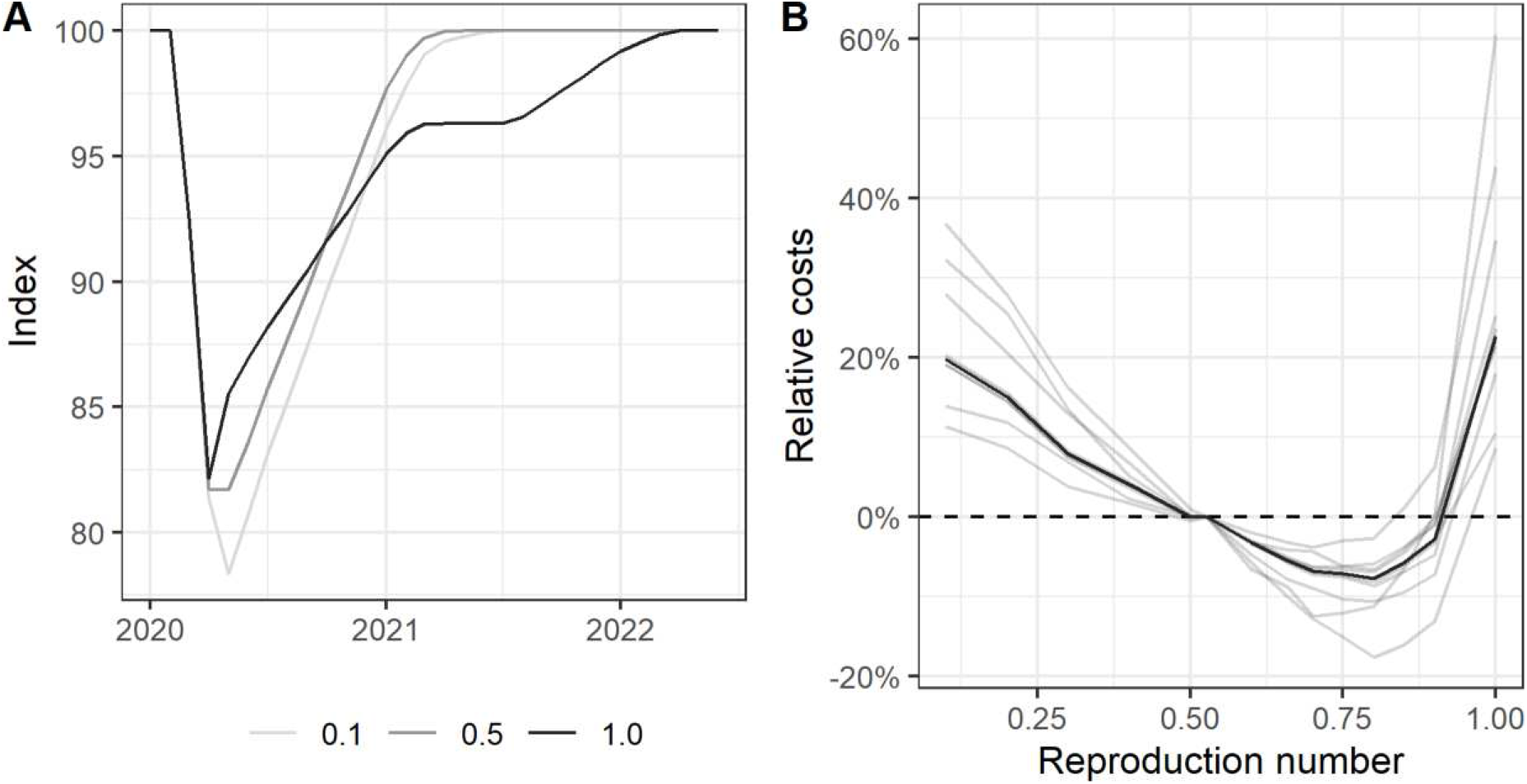
**(A)** Overall economic activity over time for three baseline policy scenarios (denoted by their respective reproduction numbers, 0.1, 0.5, and 1.0). Pre-shutdown economic activity is normalized to 100. **(B)** Relative costs for each policy scenario, in percentage difference to the reference scenario (*R*_t_ = 0.53). Economic costs are given as the aggregated loss of activity occurring as a result of the shutdown and recovery phase. The bold line indicates the baseline scenarios; the shaded grey lines indicate the results of the robustness tests. The numeric values can be found in the supplement, Table S3.

## Economy and health have a common interest of slow NPI release

Thus, we cannot identify a conflict between the economy and health protection in relation to a strong relaxation – the costs would be higher in both dimensions. Accelerated opening leads to substantially more COVID-19 deaths and increased economic costs. While strong opening policies would allow for more economic activity in the short term, our simulations suggest that the long duration of remaining restrictions would increase relative economic costs compared to alternative gradual opening strategies.

## Robustness of optimal slow NPI release suggests applicability to other countries

Our results suggest that a balanced strategy is in the common interest of health protection and the economy. The scenario calculations show that a slight, gradual lifting of shutdown restrictions is suitable to reduce the economic losses without jeopardizing medical objectives. Clearly, generalization of our results beyond Germany is limited to comparable situations and regions. The relationship between economic activity and the reproduction number might not be the same across world regions. Moreover, the shutdown duration and final death toll are influenced by the number of new infections at the point of entering or changing shutdown measures. However, our results are robust to several sensitivity tests in assumptions regarding the relationship between the shutdown severity and economic activity, affected industries of exogenous shutdown restrictions, the duration of economic recovery, and the number of daily new cases that needs to be reached to control the epidemic. Moreover, we tested the sensitivity of our assumption on the time of large scale availability of a vaccine. The robustness tests can be found in the supplement. Our inferences do not change. The economically optimal strategy is at a reproduction number around 0.8 in all sensitivity tests (see light grey lines in Fig. 4B).

We consider the qualitative statement of our results to be robust and of general nature. Our findings support a prudent, gradual opening strategy. This result is in line with retrospective studies of the influenza pandemic in 1918 in the USA^8,18^. It is in common interest of health and the economy to implement opening policies with prudent steps and to closely monitor the respective reaction of the infection figures. Using counteracting measures – such as face masks, behavioral rules or increased testing – may limit the spread of the virus^19,20^ during opening and creates leeway for larger opening and economic recovery. The level of economic restrictions thus depends to a large extent on behavioral adjustments of the population to learn to live with the virus until a vaccine or effective medical treatment is available.

## Data Availability

Not applicable

## Authors’ contribution

FD, MS, TW, and SL developed the economic model. The model is based on earlier work by TW that was extended for the purposes of this study by MS, with support from FD, TW, SL, AP, and CF. MS was responsible for the empirical implementation and simulating the policy scenarios. TW and SL were responsible for estimating economic activity on the basis of the ifo Business Survey, with support from FD, MS, and AP. FD and MS took the lead in writing the economics part of the manuscript, with support from TW, SL, AP, and CF. FD provided the literature review with support from all authors.

PV organized and prepared the raw data for the epidemiological model. BL provided evidence synthesis and epidemiological input for the epidemiological model and scenarios described here. SK, SB, and MMH designed the epidemiological scenarios, and SK performed the simulations. SK, SB, and MMH interpreted and wrote the epidemiological results.

MMH and CF designed and supervised the research. All authors contributed to the analysis and interpretation of the results. FD and MMH took the lead in writing the draft of the report (main text). The manuscript was revised by all authors. MS and SK prepared the final figures and tables.

## Acknowledgements

SB, MMH, BL and PV received funding from the European Union’s Horizon 2020 research and innovation program under grant agreement No 101003480. BL and PV were funded by the Initiative and Networking Fund of the Helmholtz Association. SB and MMH were supported by the German Federal Ministry of Education and Research within the Rapid Response Module of the National Research Network on Zoonotic Infections, project CoViDec, FKZ: 01KI20102. SK was supported by the German Federal Ministry of Education and Research within the Measures for the Establishment of Systems Medicine, project SYSIMIT (BMBF eMed project SYSIMIT, FKZ: 01ZX1308B) and by the Helmholtz Association, Zukunftsthema Immunology and Inflammation (ZT-0027).

## Supplementary Text

### Background: Containment policy in Germany

In order to contain the spread of the COVID-19 epidemic in Germany, the federal and state (Länder) governments introduced restrictive measures in several stages in March 2020. These include the banning of major events, the closure of schools and day-care centres, the forced shutdown of numerous companies in the retail, catering and many (social) services sectors, and the introduction of mobility and contact restrictions (with more or less strict curfews). In addition, many companies reduced their business activities in order to protect their employees or due to reduced demand. Citizens also adapted their behavior due to the new risk situation and information policies. The sum of these measures and behavioral changes influenced the reproduction rate in Germany: *R*_t_ fell well below 1 and the number of registered new infections per day decreased during shutdown (see Fig. S2). At the same time, however, the consequences of the COVID-19 pandemic and the shutdown measures have plunged the German economy into what is by far the deepest recession in its post-war history (Wollmershäuser et al., 2020).^1^ Since the German containment strategy seems to have been successful and the feared scenarios of casualties in Germany have so far failed to materialise, a growing public community called for faster loosening of restrictions in all areas. On April 20^th^, 2020, a conference of the federal and state governments agreed on a gradual, step-by-step loosening of the shutdown measures.^2^

### Related literature

Some macroeconomic studies address the trade-off debate between health protection and the economy. Eichenbaum et al. (2020) emphasize that the optimal containment policy saved thousands of lives, but increased the severity of the economic recession because infected people would not fully understand the effect of their decisions on the spread of the virus (Eichenbaum et al., 2020). However, an unregulated spread of the virus with larger disease burden and mortality would also have adverse effects on the economy (Correia et al., 2020; Karlsson et al., 2014). Several scholars have shown medium- and long-run economic consequences of pandemics in history (Barro et al., 2020; Jordà et al., 2020; Ma et al., 2020).

Scholars examine the optimal non-pharmaceutical intervention (NPI) strategy with respect to epidemiological and economic consequences of pandemics and conclude that (partial) shutdowns seem to be an optimal strategy (Correia et al., 2020; Holtemöller, 2020). Correia et al. (2020) examine mortality outcomes and economic effects of the 1918 “Spanish” Flu pandemic in the U.S. and find that more exposed areas by the flu experience larger and persistent declines in economic activity (Correia et al., 2020). However, by exploiting the geographical variation their results suggest that U.S. cities that intervened earlier and more aggressively by NPIs perform better in both, lower mortality rates and less adverse long-term economic consequences. In a similar vein, Holtemöller (2020) employs an integrated model assessing epidemiological and economic dynamics and concludes that mitigating the spread of the SARS-CoV-2 virus was welfare enhancing (Holtemöller, 2020). A temporary “partial” shutdown of the economy together with an extended period of intensive testing turned out to be the optimal strategy with respect to the minimal number of deaths, minimal output loss and maximal welfare. Ma et al. (2020) examine medium- and long-run economic effects of epidemics and suggest that adverse consequences were less severe in countries with larger 1^st^-year government expenditure and public health spending (Ma et al., 2020).

Several studies show that counteracting NPI measures may limit the spread of the virus and high death tolls during opening activities: Prather, Wang and Schooley (2020) as well as Mitze et al. (2020), for example, find that face masks considerably reduce the transmission of SARSCov-2 (Mitze et al., 2020; Prather et al., 2020). Grimm et al. (2020) show that a combination of tailored mechanisms, e.g. the protection of vulnerable groups together with a “trace and isolate” approach, can be effective in preventing a high death toll (Grimm et al., 2020).

The development of infection dynamics has been addressed by a number of mathematical models based on differential equation (Prem et al., 2020) and agent-based (Hoertel et al., 2020) descriptions. Similarly, economic models estimate economic costs of different scenarios (Dorn et al., 2020; IMF, 2020). However, the question of an optimal opening strategy out of a shutdown has not been addressed yet. We use empirical evidence from Germany and a novel combined epidemiological and economic model to simulate economic costs and death tolls for dynamics shaped by different scenarios of opening processes.

## Materials and Methods

### Mathematical-epidemiological model

In the SECIR model, different stages of the infection and associated viral spreading are considered. The model comprises of compartments representing the individuals susceptible (*S*) or exposed (*E*) to the virus, asymptomatic carriers (*C*_I,R_) which may become symptomatic (*I*_H,R,X_), hospitalized (*H*_U,R_) or in the need of intensive care *U*_R,D_. Infected individuals have terminal fate of recovery (*R*_Z,X_) or death (*D*). A schematic representation of the model is shown in Fig. S1A and formulated by ordinary differential equations (ODEs). The model equations read

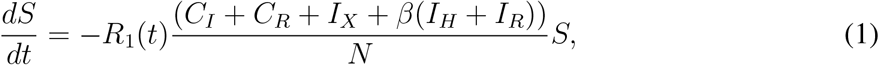

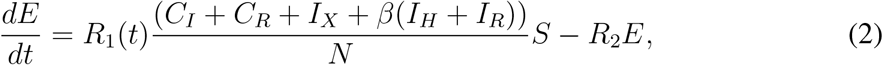

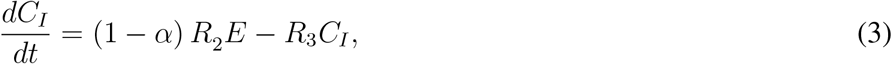

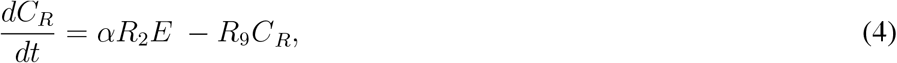

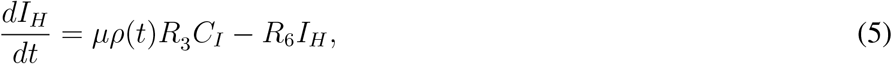

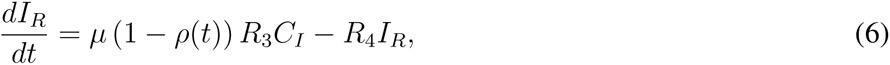

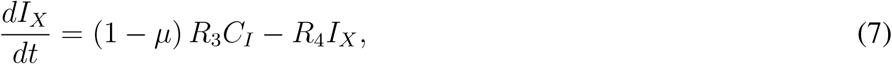

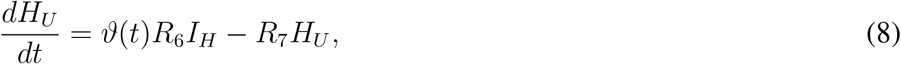

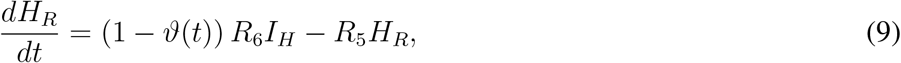

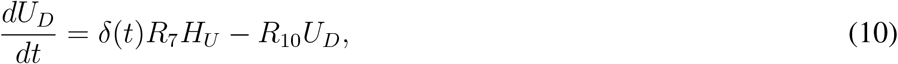

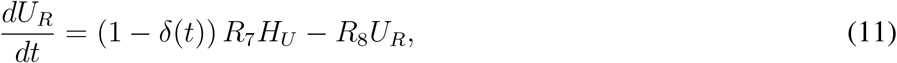

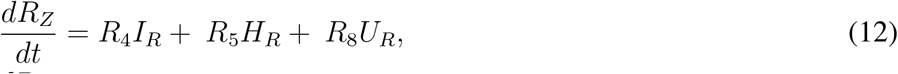

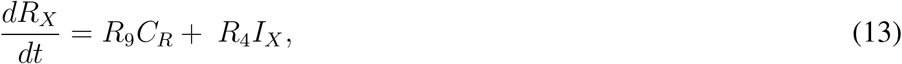

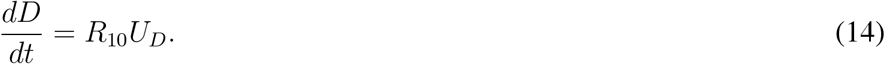

The rates R_2,…,10_ denote the inverse time of transition between the respective states and were inferred from literature. Parameter *R*_1_ is fitted to the course of reported case numbers in a sliding time window and, therefore, is a time-varying parameter. Greek letters denote fractions of individuals with a particular fate. Parameters ρ, ϑ and δ have a time-varying component modelled with a logistic function

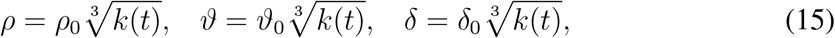

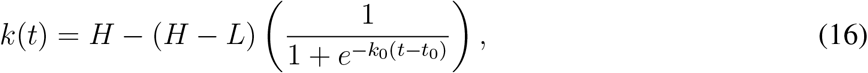

where *t* corresponds to the time after the starting of the epidemic and *H* = 0.156, *L* = 0.011, *k*_0_ = 0.25, and *t*_0_ = 61 are obtained from fitting the case fatality rate (CFR = ρϑδ) which changed over the course of the epidemic in Germany. This is due to changing testing frequencies (Robert Koch Institute, 2020) and the shifting age structure of the infected over time (Vanella et al., 2020); therefore, the CFR estimate is not reflecting a real change in the fatality rate of the virus. The uncertainty in the parameter values was incorporated into the analysis by repeated (100 times) random sampling within the plausible ranges and obtaining the distributions of model variables.

The temporal evolution of *R*_t_ was obtained by re-fitting the model in a sliding time-window of the data which was shifted throughout the duration of the epidemic (see Fig. S1B). This method allows to adapt the model parameter *R*_1_ that is associated with NPIs in order to fit the data, which cannot be achieved by a fixed parameter value. The value of *R*_t_ corresponding to *k*-th time-window is calculated based on the fitted value of *R*_1_ by

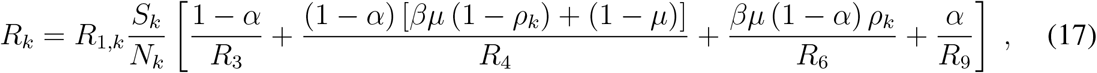

where *ρ_k_*: = *ρ*(*t_k_*) denotes the average value of the time-varying parameter in the *k*-th time-window.

For simulating prospective dynamics of the epidemic from a defined starting date, the state condition of the model based on available retrospective data was calculated. Then, the development of model variables was obtained by imposing fixed *R*_1_ values corresponding to different *R*_t_s of interest (see Fig. S1C). The assumed prospective *R*_t_s values can be linked to the different strictness of the imposed measures.

**Table S1:**
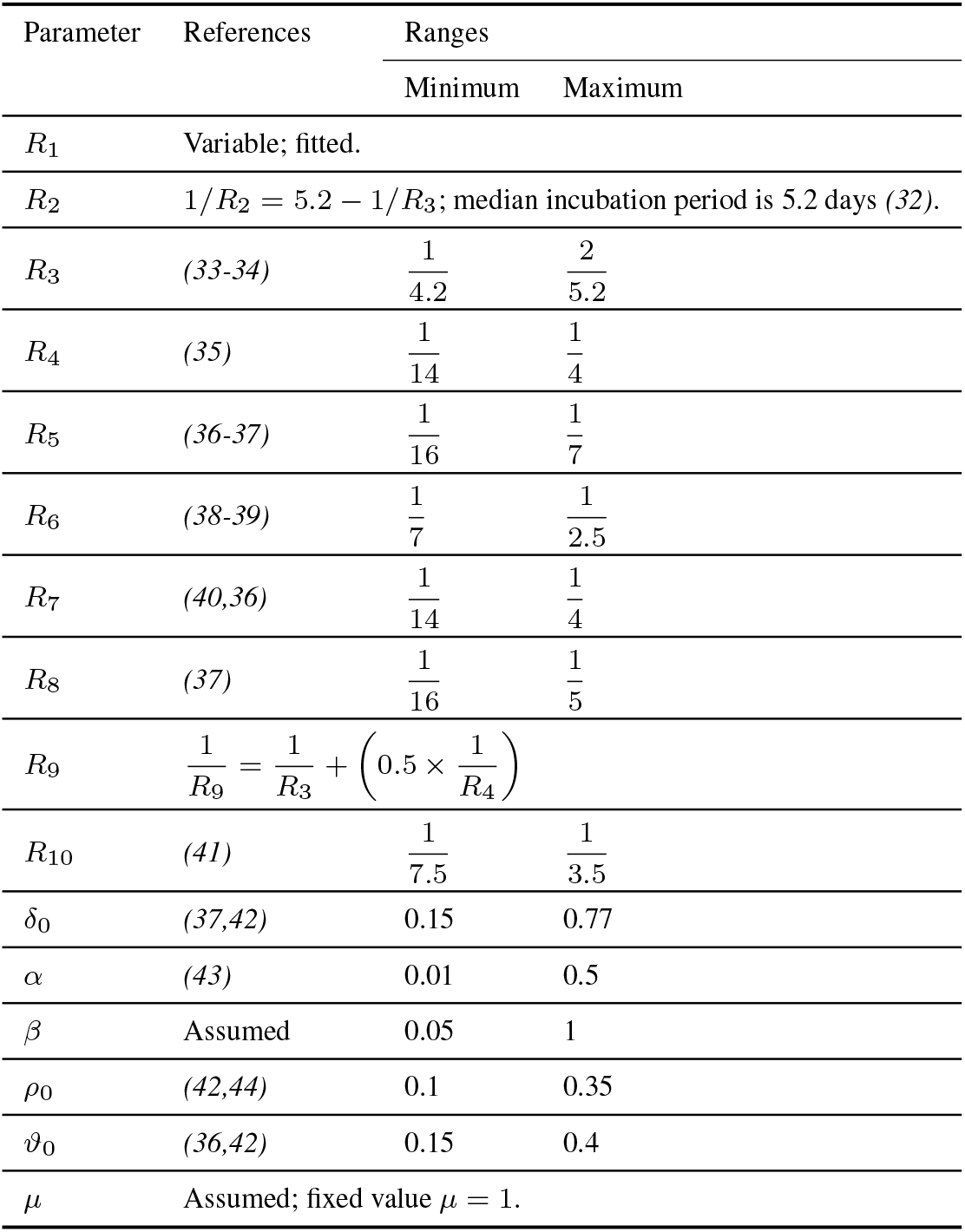
Parameter sets of the SECIR model.

| Parameter | References | Ranges |  |
| --- | --- | --- | --- |
|  |  | Minimum | Maximum |
| $R_1$ | Variable; fitted. | | |
| $R_2$ | $1/R_2 = 5.2 - 1/R_3$; median incubation period is 5.2 days (32). | | |
| $R_3$ | (33-34) | $\frac{1}{4.2}$ | $\frac{2}{5.2}$ |
| $R_4$ | (35) | $\frac{1}{14}$ | $\frac{1}{4}$ |
| $R_5$ | (36-37) | $\frac{1}{16}$ | $\frac{1}{7}$ |
| $R_6$ | (38-39) | $\frac{1}{7}$ | $\frac{1}{2.5}$ |
| $R_7$ | (40,36) | $\frac{1}{14}$ | $\frac{1}{4}$ |
| $R_8$ | (37) | $\frac{1}{16}$ | $\frac{1}{5}$ |
| $R_9$ | $\frac{1}{R_9} = \frac{1}{R_3} + \left(0.5 \times \frac{1}{R_4}\right)$ | | |
| $R_{10}$ | (41) | $\frac{1}{7.5}$ | $\frac{1}{3.5}$ |
| $\delta_0$ | (37,42) | 0.15 | 0.77 |
| $\alpha$ | (43) | 0.01 | 0.5 |
| $\beta$ | Assumed | 0.05 | 1 |
| $\rho_0$ | (42,44) | 0.1 | 0.35 |
| $\vartheta_0$ | (36,42) | 0.15 | 0.4 |
| $\mu$ | Assumed; fixed value $\mu = 1$. | | |

### Time until new infections are under control

We assume that the approximately 400 health authorities in Germany have sufficient capacities to control 300 new cases per day through contact tracing and isolation. For the bundles of measures with varying severity, we calculated the time it takes to reach a maximum of 300 new reported cases per day (Fig. 2B) and the number of projected COVID-19 deaths (Fig. 2A). This was calculated by obtaining the daily influx of cases to the infected and recovering components *I*_H_ and *I*_R_. The analysis was also repeated for capacities of 200 and 400 new cases per day in our robustness tests to reflect the sensitivity of the impact of different assumptions in the capacity of health authorities. Once the respective assumed threshold is reached, an *R*_t_ value of 1 and a targeted isolation of identified newly infected and their contacts (β = 0) were assumed. These assumptions keep the number of new infections around the target value. Alternatively, we have considered a scenario in which the current daily infection rates are kept constant until the earliest realistic availability date of a vaccine for the entire population, i.e. *R*_t_ is assumed to be at value of 1.

The calculated duration shall be interpreted as the further time necessary to keep the shutdown or restrictive measures. Based on the reduction in economic output in the various economic sectors on April 20^th^, 2020, and the *R*_t_ value corresponding to the shutdown period (i.e. in the status quo before the first relaxation of measures on April 20^th^, 2020), the costs of maintaining the shutdown or restrictive measures until reaching 300 (or 200 or 400) new reported cases per day were estimated for each assumed scenario with *R*_t_ < 1. For the scenario of *R*_t_ = 1, we assume that there will be restrictive measures until a vaccine becomes generally available. The Paul Ehrlich Institute^3^ estimates that combined Phase II/III trials of a vaccine could start in autumn/winter 2020. For our scenario we therefore assumed this date to be July 31^st^, 2021. In our sensitivity tests, we assume the vaccine to be available at a large scale 120 days earlier or later compared to the baseline assumption.

### NPIs and their impact on *R*_t_

In order to quantify the impact of lifting measures, we assumed a 2-weeks time delay from a person being exposed to the virus to become symptomatic and reported in the database. For the first openings in Germany on April 20^th^, 2020, the reporting delay assumption implies that a person exposed on the first day of lifting measures will be reported on May 4^th^. However, as the *R*_t_ value reported on each date includes the impact of 6 other days in the data retrospectively (see Fig. S1B), the *R*_t_ value calculated on May 4^th^is biased by cases exposed before April 20^th^(6 out of 7 data-points). With a similar reasoning, the *R*_t_ values calculated in the period of May 4^th^-May 9^th^are contaminated with infected cases before April 20^th^, 2020. Therefore, the impact of the openings on April 20^th^shall be inferred from May 10^th^at the earliest. Since the obligation of wearing masks was imposed very shortly after the first openings, we considered them as a bundle of NPIs. Following the reporting delay assumption, the impact of the NPIs bundle on *R*_t_ value is expected from May 19^th^at the earliest. In order to take into account the seasonality observed in the data and the *R*_t_ values in the calculation of NPI impacts, we considered a pooled set of *R*_t_ values in the 1-week period of May 19^th^-May 25^th^(see Fig. S2 and S3).

**Figure S1:**
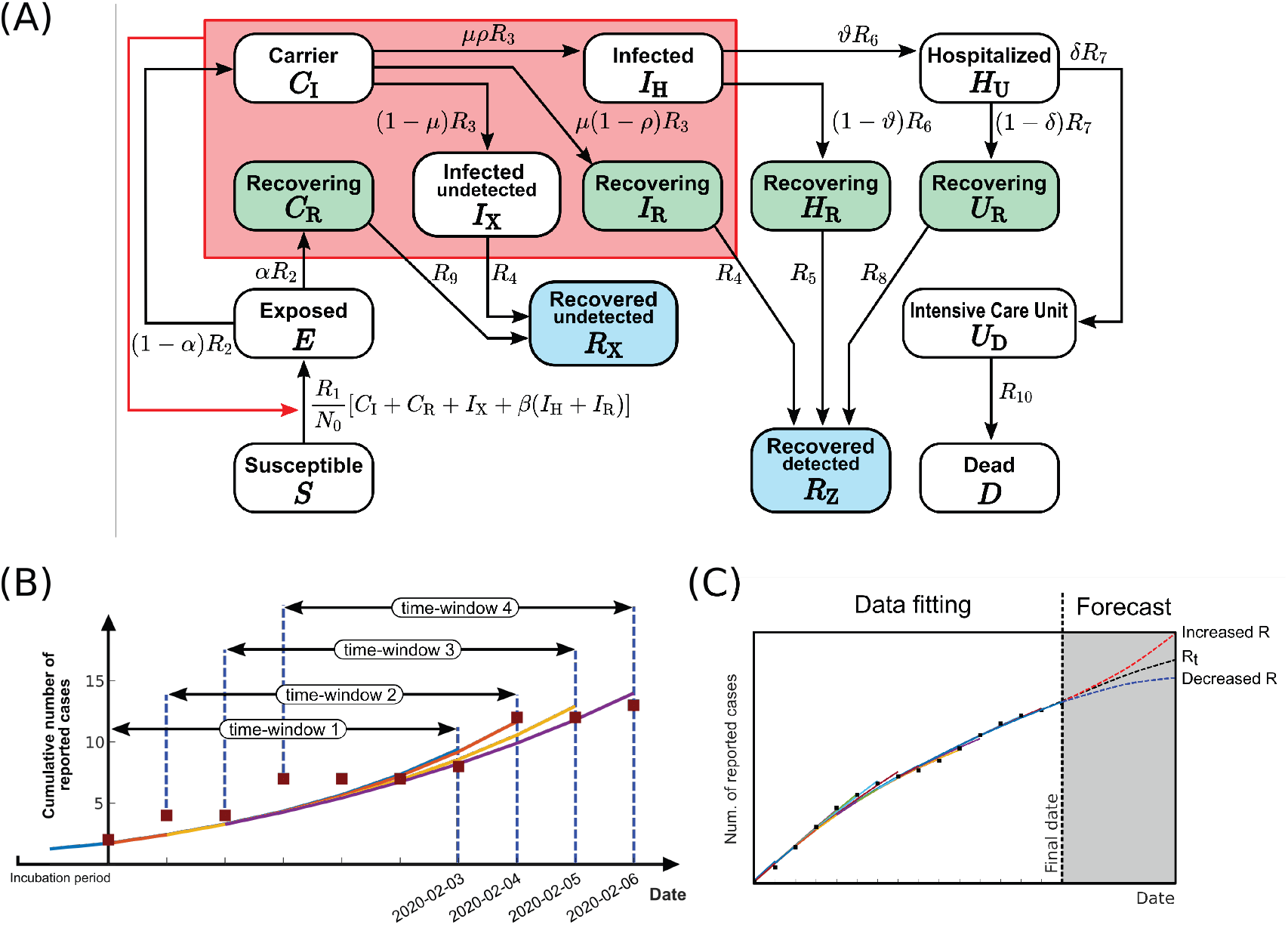
(A) The scheme of the SECIR model. The model distinguishes susceptible (*S*), healthy individuals without immune memory of CoV, exposed (*E*), who carry the virus but are not yet infectious to others, carriers (*C*_I,R_), who carry the virus and are infectious to others but do not yet show symptoms, infected (*I*_H,R,X_), who carry the virus with symptoms and are infectious to others, hospitalized (*H*_U,R_), who experience a severe development of the disease, transferred to intensive care unit (*U*_R,D_), dead (*D*), and recovered (*R*_D,X_), who acquired immune memory and cannot be infected again. Recovery happens from each of the states *C*_R_, *I*_X_, *I*_R_, *H*_R_, *U*_R_. See Table S1 for parameter values. (B) Algorithm of calculating time-varying reproduction number with sliding time-window. Starting from an exposed population based on the initial case reports, the parameter *R*_1_ was fitted to the 1-week time window in the data and the corresponding *R*_t_ value was calculated. Next, starting from the state condition of the model at the first time-point, the fitting process was repeated for the time-window shifted by 1 day. This process was repeated for the whole duration of the epidemic. The calculated *R*_t_ value was reported for the final date of each time-window. (C) Scheme of prospective simulations. Time evolution of the model variables was obtained from the case reports until the starting date of the prospective study. Then, starting from the last state condition of the model, the numerical simulation was continued with imposed fixed values of R_1_ that correspond to the *R*_t_ values of interest.

**Figure S2:**
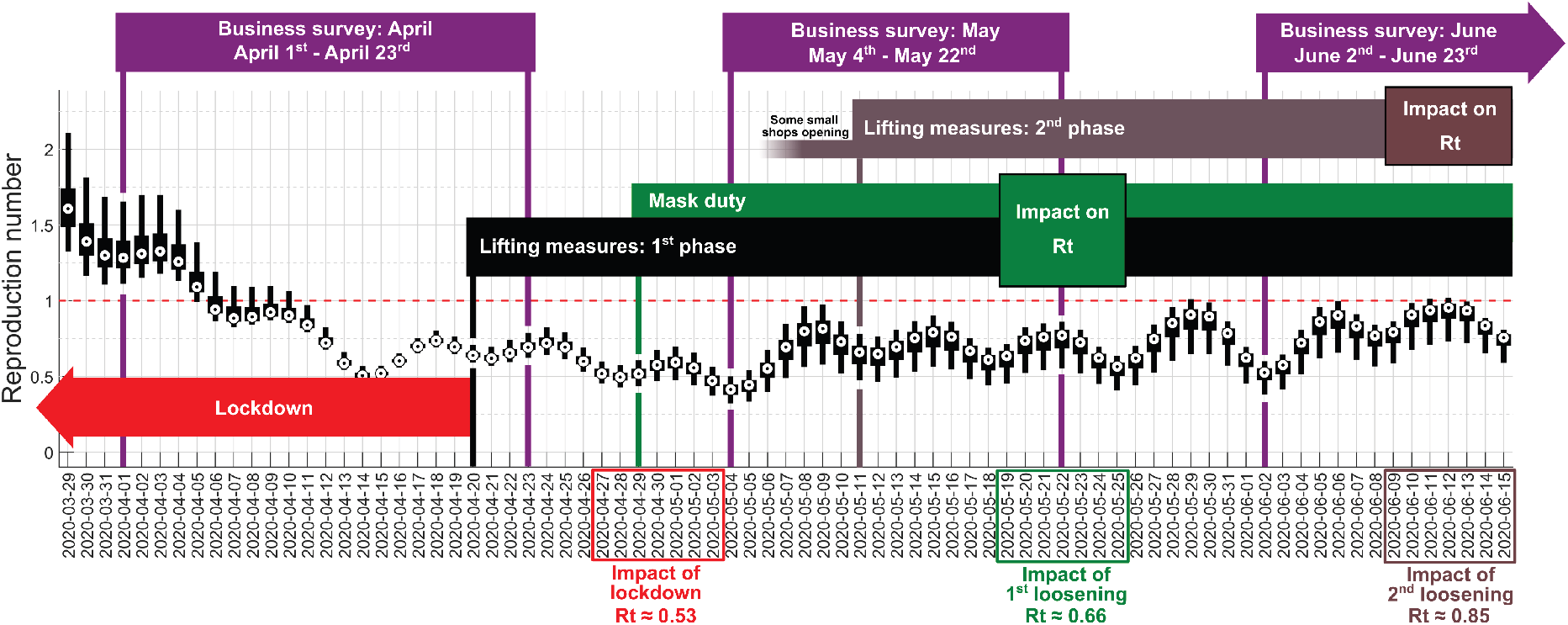
Time evolution of reproduction number in Germany. The timeline of business surveys and NPIs are marked. The time-windows used for pooling *R*_t_ values associated with each NPI is shown on the horizontal axis. The boxplots illustrate the median, 25- and 75-percentiles, maximum and minimum values.

**Figure S3:**
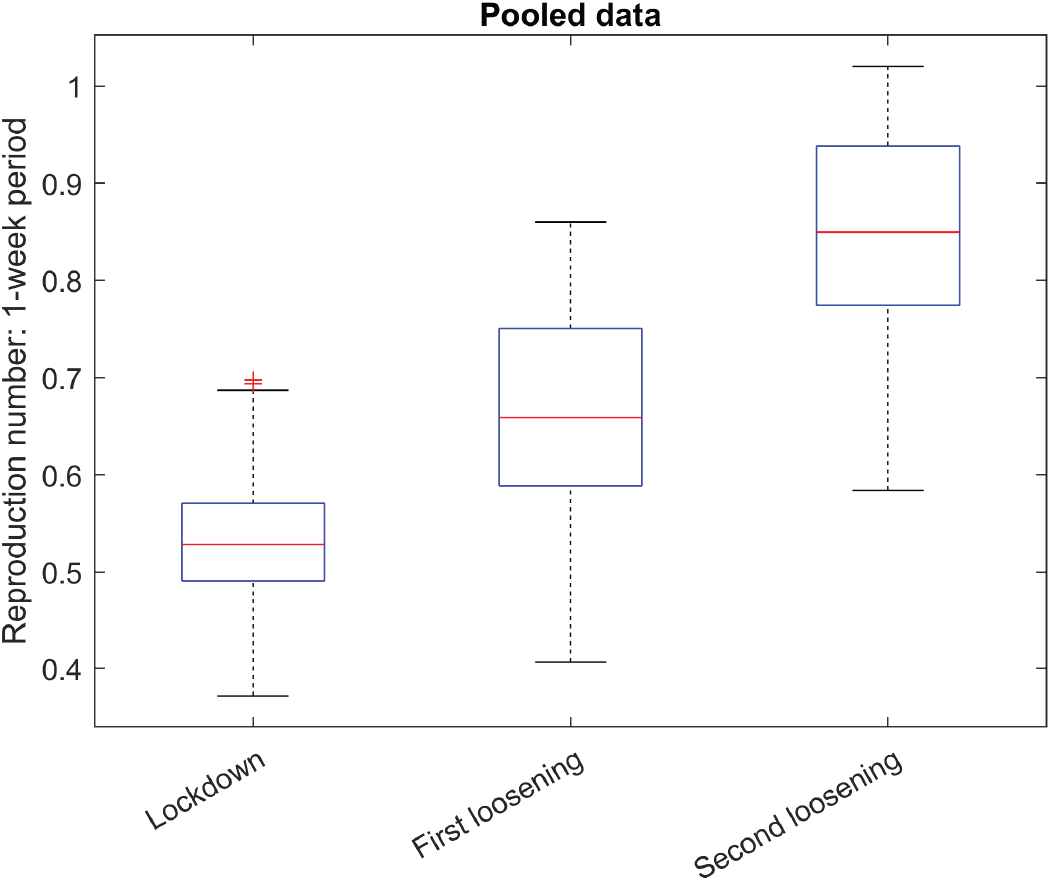
Distribution of *R*_t_ values associated with NPIs. Each scenario and corresponding time-window for pooling the data are shown in Fig. S2. The boxplots illustrate the median, 25-and 75-percentiles, maximum and minimum values. The median was used for the economical model.

The *R*_t_ value corresponding to the complete shutdown (before April 20^th^) was calculated by excluding the contaminated period (May 4^th^-May 9^th^), following a similar reasoning. Herefore we considered the *R*_t_ values at the latest possible week, the period of April 27^th^-May 3^rd^. The impact of the second nationwide lifting of measures was calculated by pooling the latest available week at the time of this analysis (see Fig. S2 and S3).

## Economic model and empirical implementation

### Modelling economic costs

The economic costs of scenario *s* are given as the aggregated loss of activity occurring as a result of the shutdown. Denote 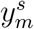 as the economic activity compared to the pre-shutdown level in scenario *s* and month *m*, with 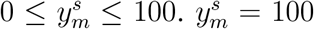 refers to the pre-shutdown activity level, and 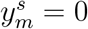 to an economy with zero production. Total costs of scenario s can be written as:

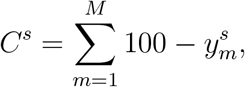

where *M* is the time horizon under consideration, i.e. the total number of months that are taken into account in the analysis. Denote *C^ref^* as the cost of a reference scenario. The relative costs of *s* are then given as ∆*C^s^* = (*C^s^/C^ref^*) − 1, such that ∆*C^s^* > 0, indicating scenarios with higher costs and lower aggregate economic activity compared to the reference.

The key challenge is to model 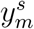. We assume that 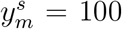 prior to the implementation of the measures, 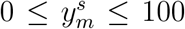 during the shutdown and the recovery phase, and 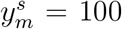 again after the recovery phase. In other words, starting from the pre-shutdown activity level, activity drops during the shutdown, and recovers once the measures are lifted until the economy has returned to its activity level, prior to the measures.

Taking into account that the impact of the shutdown varies across industries, we explicitly model activity at the industry level. Denote 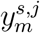 as the activity for scenario *s*, month *m*, and industry 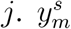 is then given as the average of each industry-specific activity, weighted by the share of the industry in total output, denoted as *α_j_*:

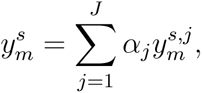

where *J* is the total number of industries in the economy.

For each *s* and *j*, the process of 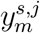 over *m* is modelled as follows. Denote 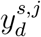 as the activity on day *d*, with 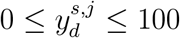 and *d* ∈ {1,…, *D*}, where *D* is the last calendar day in the last year of observation. 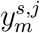 is given as

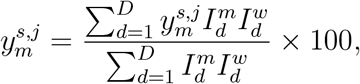

where 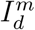 is an indicator variable equal to one if calendar day d belongs to calendar month *m*, and zero otherwise. Similarly, 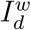 indicates whether the calendar day is a working day or not (i.e. whether it falls on a weekend or a public holiday). This notation implies that the distribution of holidays across the calendar year is relevant for the cost of a scenario. That is, the shutdown is less costly when it is in place during months with few working days.

Further, denote *B* as the calendar day when shutdown measures were implemented first, S the day when a new policy is introduced (changing the severity of the measures), and *R^s,j^* the day when the measures are lifted. *R^s,j^* is determined by the epidemiological model and describes the calendar day during which a certain daily case number has been reached. The superscripts indicate that there is heterogeneity across scenarios and industries. After the introduction of the new policy at *S* the economy adjusts over the period *s^s,j^*, after which the new activity level is reached. After the prescribed number of new infections is reached and the shutdown is fully lifted, i.e. for *d* > R, the economy slowly recovers and returns to pre-shutdown activity. We assume that economic activity increases linearly from *S* to *S* + *s^s,j^* and from *R^s,j^* to *R^s,j^*+ *r^s,j^*. *r^s,j^* denotes the industry-specific duration of the recovery period (in calendar days). For each day d, the activity in each scenario *s* and sector *j* is then given as follows:

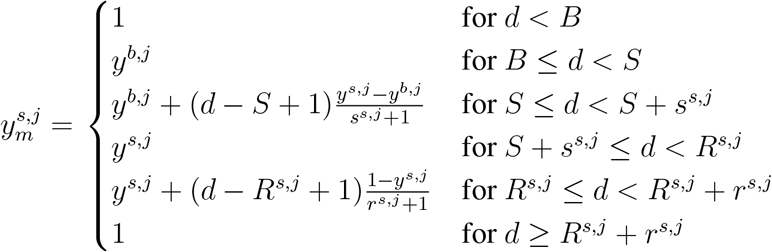

*y^b,j^* and *y^s,j^* refer to the activity after the first introduction of shutdown measures and after the policy change, respectively. Figure S4 illustrates the process of 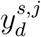 over *d*.

**Figure S4:**
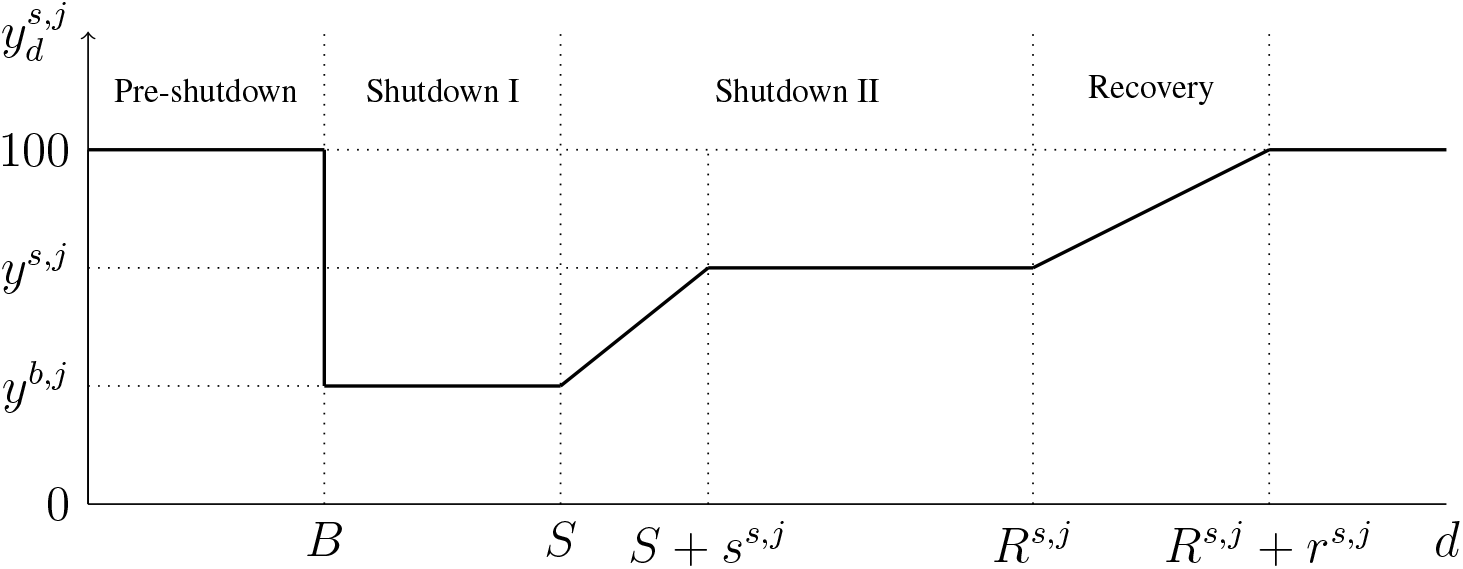
The figure illustrates the process of economic activity in the model. Starting from a pre-shutdown level, the economy experiences a decline in activity during the shutdown (from 100 to *y^b,j^*). While the measures are in place, the policy-makers may adjust their severity (from *y^b,j^* to *y^s,j^*). During the recovery phase, the economy slowly returns to its pre-shutdown level (from *y^s,j^* to 100).

### Implementing the economic model

Implementing the economic model requires estimates for *y^b,j^*, *y^s,j^*, *R^s,j^*, and *r^s,j^*. *B, S*, and *s^s,j^* are determined exogenously. For our application, we specify March 19^th^, 2020 as the introduction date of the shutdown measures (i.e. *B* = 79). *S* refers to April 20^th^, when the federal and state governments agreed on a gradual, step-by-step loosening of the shutdown measures (i.e. *S* = 111). *s^s,j^* is set to 21. That is, we assume that it takes an industry three weeks to adjust and reach the new activity level. The specification of *s^s,j^* is arguably ad-hoc, but the results are robust to different specifications. *R^s,j^* is estimated with the SECIR model. In general, a higher (smaller) *R*_t_ value is associated with a higher (smaller) *R^s,j^*.

Estimates for *y^b,j^*, *y^s,j^*, and *r^s,j^*are obtained on the basis of the ifo Business Survey, a long-running monthly panel survey of roughly 9,000 German firms (Sauer & Wohlrabe, 2020). *y^b,j^* and *y^s,j^*are approximated by the companies’ assessment of the current business situation, which they can describe as “good”, “satisfactory”, or “poor”. For each economic section of the NACE Rev. 2 classification (except for sections A, B, D, and E, as well as O, P, and Q) the responses are summarized as balance statistics, which are calculated as the difference in the percentage shares of the responses “good” and “poor”. For both, the aggregate economy and specific industries, these balance statistics are highly correlated with gross value added and other official activity measures provided by the Federal Statistical Office of Germany (Lehmann, 2020).

While *y^b,j^*is constant across all scenarios, *y^s,j^* is allowed to vary. We start by normalizing the activity level of the model prior the shutdown on March 19^th^ to zero. The activity prior to the shutdown refers to the average of the balance statistics of the business situation in January and February 2020. We refer to this as the baseline business situation. To obtain an estimate of *y^b,j^*, for each industry we first compute the difference between the balance statistic of the business situation during the shutdown in April 2020 and the baseline business situation (see Table S2). We then apply the following transformation to the balance point differences, ensuring that 0 ≤ *y^b,j^*≤ 100:

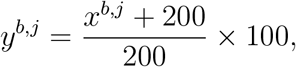

where *x^b,j^* are the balance points differences. Note that the balance point differences are not meant to reflect absolute differences of gross value added. Instead, they indicate the relative degree to which industries are hit during the shutdown and how they perform thereafter. The transformation such that *y^b,j^* is between 0 and 100 captures the intuition that economic capacity can range from zero to full capacity.

The activity level after the introduction of the new policy, *y^s,j^*, is estimated in several steps. We first calculate the difference between the balance statistic of the business situation in June 2020 and the baseline business situation (see Table S2), and again apply the transformation described above. This yields the change in economic activity that is associated with the gradual lifting of the shutdown measured in Germany after April 20^th^. From the SECIR model we obtain two corresponding *R*_t_ values, one referring to the reproduction number before (*R*_t_ = 0.53) and one after the lifting (*R*_t_ = 0.85). To obtain estimates for *y^s,j^* for all values of *R*_t_, we assume the relationships between the observed change in *R*_t_ and the industry-specific changes in economic activity to be linear. For instance, in a scenario where we simulate an increase of *R*_t_ that is twice as much compared to the observed change from 0.53 to 0.85, economic activity in each industry also increases twice as much.

Note that in the case of Germany, not all industries were affected exogenously by the shutdown measures in the sense that the measures were imposed by the government. For instance, the shutdown of businesses in mid-March did not apply to manufacturing firms. Still, we observe a drop in production in these firms. However, this is rather due to an endogenous reaction to sluggish demand, disrupted supply chains, or a shortage in labor supply. We take this into account and distinguish between exogenous and endogenous industries (see Table S2). For *exogenous* industries, *y^s,j^*is estimated as described above. In contrast, for *endogenous* industries, we calculate the activity level based on an input-output matrix, which specifies to what extent the production in one industry is affected by changes in production in another industry (Federal Statistical Office, 2020). We use the input-output matrix to calculate the change in activity level for each endogenous industry, ∆_*yi*_, based on the changes in activity levels in all exogenous industries:

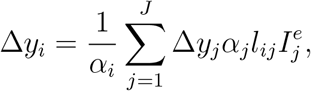

for *i* ≠ *j*. *l_ij_* specifies the change in output in industry *i* that is due to a unit change in output in industry *j*. *α_i_* and *α_j_* are the the respective industry’s share in total economic output. 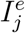 is an indicator variable equal to one if industry *j* is an exogenous industry, and zero otherwise. We additionally assume the shutdown duration to be constant across all endogenous industries and scenarios. Specifically, we set the the shutdown duration *R^s,j^* − *S* = 30, i.e. the duration in the reference scenario.

Finally, the estimate for *r^s,j^*, the duration of the recovery period of industry *j* in scenario *s*, is based on a special question in the ifo Business Survey in May. Respondents were asked about the expected duration until their business situation would return to normal once the shutdown measures are lifted. For the reference scenario (*R*_t_ = 0.53), we take the mean of these expectations for each industry (see Table S2). Similar to *y^s,j^*, all other scenarios assume a linear relationship between *R*_t_ and *r^s,j^*. To estimate the linear relationship between *R*_t_ and the recovery time, we assume that in the scenario with *R*_t_ = 0.85 it takes the firms two months less to fully recover. For the endogenous industries, we assume the recovery periods to be constant across scenarios (but not across industries) and set the recovery duration equal to the durations in the reference scenario.

**Table S2:**
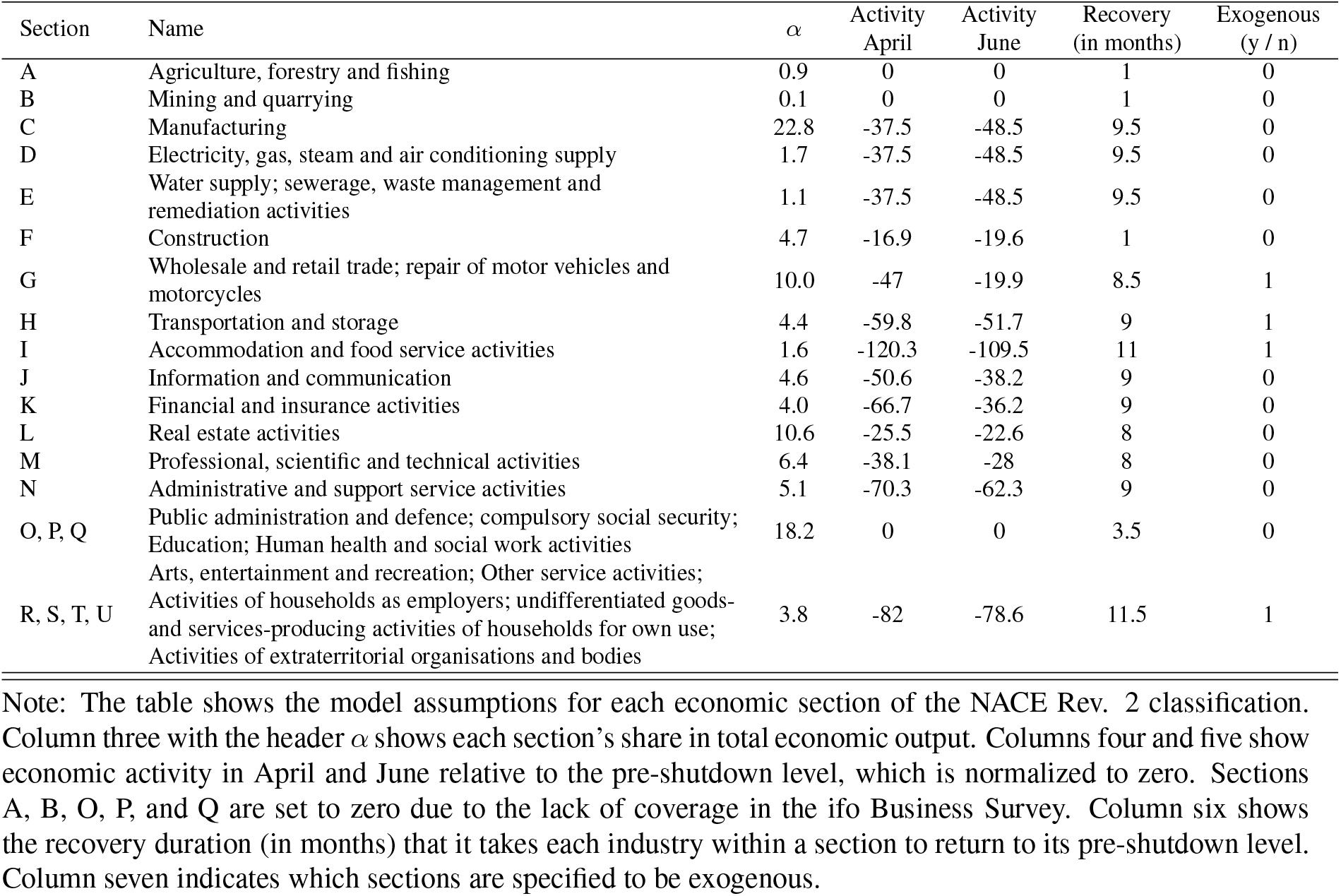
Model Assumptions

## Robustness tests

To evaluate the the robustness of our baseline result, we run a battery of sensitivity tests where we individually vary each model parameter. The results are shown in Table S3. Overall, we find that the baseline result is highly robust, confirming our main finding. In all robustness tests, costs are lowest in the scenarios with slight, step-wise loosening. The minima are all between *R*_t_ values of 0.7 and 0.8. Thus, from an economic point of view, a tightening as well as a too strong loosening of the shutdown measures is not the optimal strategy.

**Table S3:** Robustness Tests

| $R_t$ | Relative costs | | | | | | | | | | | |
| --- | --- | --- | --- | --- | --- | --- | --- | --- | --- | --- | --- | --- |
|  | Baseline | Robustness tests |  |  |  |  |  |  |  |  |  |  |
|  |  | 1 | 2 | 3 | 4 | 5 | 6 | 7 | 8 | 9 | 10 | 11 |
| 0.10 | 19.8 | 36.7 | 11.3 | 13.9 | 27.9 | 19.0 | 20.3 | 20.0 | 19.2 | 32.3 | 19.8 | 19.8 |
| 0.20 | 15.0 | 27.8 | 8.6 | 11.8 | 20.6 | 14.4 | 15.5 | 15.2 | 14.6 | 25.5 | 15.0 | 15.0 |
| 0.30 | 7.9 | 16.2 | 3.7 | 6.9 | 13.0 | 7.6 | 8.2 | 7.9 | 7.5 | 13.4 | 7.9 | 7.9 |
| 0.40 | 4.0 | 8.7 | 1.7 | 2.2 | 6.8 | 3.9 | 4.4 | 4.0 | 3.7 | 5.1 | 4.0 | 4.0 |
| 0.50 | -0.1 | 0.9 | -0.6 | -0.1 | -0.1 | -0.1 | -0.1 | -0.1 | -0.2 | 0.2 | -0.1 | -0.1 |
| 0.53 | 0.0 | 0.0 | 0.0 | 0.0 | 0.0 | 0.0 | 0.0 | 0.0 | 0.0 | 0.0 | 0.0 | 0.0 |
| 0.60 | -3.2 | -5.8 | -2.0 | -3.2 | -4.7 | -3.0 | -3.2 | -3.4 | -3.3 | -6.6 | -3.2 | -3.2 |
| 0.66 | -5.5 | -10.0 | -3.3 | -4.1 | -7.8 | -5.1 | -5.8 | -5.7 | -5.3 | -8.8 | -5.5 | -5.5 |
| 0.70 | -6.9 | -12.9 | **-3.8** | -4.4 | -9.1 | -6.4 | -7.2 | -7.0 | -6.5 | **-12.5** | -6.9 | -6.9 |
| 0.75 | -7.1 | -15.1 | -3.0 | **-6.2** | -10.4 | -6.2 | -7.6 | -7.4 | -6.6 | -12.1 | -7.1 | -7.1 |
| 0.80 | **-7.8** | **-17.7** | -2.7 | -5.9 | **-10.7** | **-6.7** | **-8.7** | **-8.2** | **-7.0** | -11.3 | **-7.8** | **-7.8** |
| 0.85 | -5.8 | -16.1 | 1.0 | -3.9 | -9.5 | -4.2 | -6.9 | -6.3 | -4.5 | -6.6 | -5.8 | -5.8 |
| 0.90 | -2.8 | -13.1 | 6.2 | -1.0 | -7.3 | -0.1 | -4.8 | -3.3 | -0.8 | 0.3 | -2.8 | -2.8 |
| 1.00 | 22.6 | 8.7 | 43.9 | 25.2 | 18.0 | 21.4 | 23.7 | 22.8 | 22.0 | 60.4 | 10.5 | 34.7 |

Note: The table shows the relative costs for the baseline and the robustness tests. Relative costs are given as the percentage differences in total loss of economic activity compared to the reference scenario ( $R_t = 0.53$ ). For the baseline and each robustness test, the $R_t$ value associated with the lowest costs are indicated in bold.

In the first two robustness tests, we vary the linear-relationship assumption between the reproduction number and economic activity. We re-scale the estimated coefficient slope for each industry by the factors 2 and 0.5, respectively. That is, we assume that the change in activity when changing the reproduction number is double (half) in magnitude compared to the baseline. Similarly, we vary the linear-relationship assumption between the reproduction number and the duration of the recovery period. Again, we re-scale the coefficient slope by the factor 2 (0.5), i.e. it takes each industry double (half) the time to fully recover from the shutdown.

In a next robustness test, we specify all service industries (sections J to Q in Table S2) to be exogenously affected by the shutdown measures. This controls for the potential issue that the government might have more control over economic activity than we assume in the baseline.

The remaining robustness tests vary the assumptions about the shutdown duration. First, we change the number of required number of new infections per day from 300 to 200 and 400, respectively. This captures the intuition that policy-makers might aim at lower or higher daily cases numbers. Next, since the estimated duration to reach the 300 daily cases for each *R*_t_ value is subject to sampling uncertainty, we additionally calibrate our model using the 2.5 and 97.5 percentile of the distribution, i.e. assuming that it take less and more days to reach the 300 cases. And finally, we vary the period when a vaccine becomes available at large scale, by shifting the date forward and backward by 120 days compared to the baseline.

## Footnotes

1 It can be assumed that an unhindered spread of the virus would also have been associated with very high economic and health costs.

2 The actual death toll for Germany counts around 4,500 registered COVID-19 deaths on April 20^th^, around 6,500 two weeks later at the beginning of May, around 8,500 at the beginning of June, and around 9,000 at the beginning of July 2020. Germany has a population of around 83 million (Johns Hopkins University, 2020). The death toll corresponds to relative numbers of around 5.3 deaths per 100k inhabitants (April 20th), 9.7 (one month later) and 10.6 (two month later). In comparison two month after the start of relaxations, relative death rates are around seven times larger in the UK, around five times larger in Sweden, and around four times larger in the USA (Human Mortality Database, 2020a, 2020b, 2020c).

3 The Paul Ehrlich Institute is a German research institution and medical regulatory body, and is the German federal institute for vaccines and biomedicines.

## References

1 Ferguson, N. et al. Report 9: Impact of non-pharmaceutical interventions (NPIs) to reduce COVID19 mortality and healthcare demand. (Imperial College London, 2020).

2 Dehning, J. et al. Inferring change points in the spread of COVID-19 reveals the effectiveness of interventions. Science 369, eabb9789 (2020).

3 Dorn, F. et al. The economic costs of the coronavirus shutdown for selected European countries: A scenario calculation. EconPol Policy Brief 25 (2020).

4 IMF. World Economic Outlook, April 2020: The Great Lockdown. (International Monetary Fund, 2020).

5 Eichenbaum, M. S., Rebelo, S. & Trabandt, M. The macroeconomics of epidemics. NBER Working Paper 26882 (2020).

6 The Economist. A grim calculus: Covid-19 presents stark choices between life, death and the economy. The Economist, April 2nd (2020).

7 Karlsson, M., Nilsson, T. & Pichler, S. The impact of the 1918 Spanish flu epidemic on economic performance in Sweden: An investigation into the consequences of an extraordinary mortality shock. Journal of Health Economics 36, 1–19 (2014).

8 Correia, S., Luck, S. & Verner, E. Pandemics depress the economy, public health interventions do not: evidence from the 1918 Flu. SSRN (2020).

9 Barro, R. J., Ursua, J. e. F. & Weng, J. The coronavirus and the great influenza epidemic –Lessons from the “Spanish Flu” for the coronavirus’s potential effects on mortality and economic activity. CESifo Working Paper 8166 (2020).

10 Jordà, Ò., Singh, S. R. & Taylor, A. M. Longer-run economic consequences of pandemics. NBER Working Paper 26934 (2020).

11 Ma, C., Rogers, J. H. & Zhou, S. Global economic and financial effects of 21st century pandemics and epidemics. SSRN (2020).

12 Abele-Brehm, A. et al. Making the fight against the Coronavirus pandemic sustainable. (ifo Institute, 2020).

13 Holtemöller, O. Integrated assessment of epidemic and economic dynamics. IWH Discussion Papers 4/2020 (2020).

14 Khailaie, S. et al. Estimate of the development of the epidemic reproduction number Rt from Coronavirus SARS-CoV-2 case data and implications for political measures based on prognostics. medRxiv (2020).

15 Sauer, S. & Wohlrabe, K. ifo Handbuch der Konjunkturumfragen. (ifo Beiträge zur Wirtschaftsforschung, 2020).

16 Lehmann, R. The Forecasting Power of the ifo Business Survey. CESifo Working Paper 8291 (2020).

17 Federal Statistical Office. Volkswirtschaftliche Gesamtrechnungen. Input-Output-Rechnung nach 12 Gutergruppen / Wirtschafts-und Produktionsbereichen. (Federal Statistical Office of Germany, 2020).

18 Markel, H. et al. Nonpharmaceutical interventions implemented by US cities during the 1918–1919 influenza pandemic. JAMA 298, 644–654 (2007).

19 Prather, K. A., Wang, C. C. & Schooley, R. T. Reducing transmission of SARS-CoV-2. Science 368, 1422–1424 (2020).

20 Mitze, T., Kosfeld, R., Rode, J. & Wälde, Klaus. Face Masks Considerably Reduce COVID-19 Cases in Germany: A Synthetic Control Method Approach. IZA Discussion Paper 13319 (2020).

## References

Barro, R. J., Ursua, J. F., & Weng, J. (2020). The coronavirus and the great influenza epidemic – lessons from the “spanish flu” for the coronavirus’s potential effects on mortality and economic activity. CESifo Working Paper, 8166.

Correia, S., Luck, S., & Verner, E. (2020). Pandemics depress the economy, public health interventions do not: Evidence from the 1918 flu. SSRN. https://dx.doi.org/10.2139/ssrn.3561560

Dorn, F., Fuest, C., Göttert, M., Krolage, C., Lautenbacher, S., Lehmann, R., Link, S., Möhrle, S., Peichl, A., Reif, M., Et al. (2020). The economic costs of the coronavirus shutdown for selected European countries: A scenario calculation. EconPol Policy Brief, 25.

Eichenbaum, M. S., Rebelo, S., & Trabandt, M. (2020). The macroeconomics of epidemics. NBER Working Paper, 26882.

Federal Statistical Office. (2020). Volkswirtschaftliche Gesamtrechnungen. Input-Output-Rechnung nach 12 Gütergruppen / Wirtschafts-und Produktionsbereichen. Federal Statistical Office of Germany.

Grimm, V., Mengel, F., & Schmidt, M. (2020). Extensions of the SEIR model for the analysis of tailored social distancing and tracing approaches to cope with COVID-19. medRxiv. https://www.medrxiv.org/content/10.1101/2020.04.24.20078113v1

Hoertel, N., Blachier, M., Blanco, C., Olfson, M., Massetti, M., Rico, M. S., Limosin, F., & Leleu, H. (2020). A stochastic agent-based model of the SARS-CoV-2 epidemic in france. Nature Medicine, 1–5.

Holtemöller, O. (2020). Integrated assessment of epidemic and economic dynamics. IWH Discussion Papers, 4/2020.

Human Mortality Database. (2020a). Sweden, Population size (abridged). Retrieved January 9, 2020, from https://www.mortality.org/hmd/SWE/STATS/Population.txt

Human Mortality Database. (2020b). The United States of America, Population size (abridged). Retrieved October 2, 2019, from https://www.mortality.org/hmd/GBR_NP/STATS/Population.txt

Human Mortality Database. (2020c). United Kingdom, Population size (abridged). Retrieved July 11, 2020, from https://www.mortality.org/hmd/GBR_NP/STATS/Population.txt

IMF. (2020). World economic outlook, april 2020: The great lockdown. International Monetary Fund.

Johns Hopkins University. (2020). COVID-19 Dashboard by the Center for Systems Science and Engineering (CSSE) at Johns Hopkins University (JHU). Retrieved July 30, 2020, from https://gisanddata.maps.arcgis.com/apps/opsdashboard/index.html#/bda7594740fd40299423467b48e9ecf6

Jordà, Ò., Singh, S. R., & Taylor, A. M. (2020). Longer-run economic consequences of pandemics. NBER Working Paper, 26934.

Karlsson, M., Nilsson, T., & Pichler, S. (2014). The impact of the 1918 spanish flu epidemic on economic performance in Sweden: An investigation into the consequences of an extraordinary mortality shock. Journal of Health Economics, 36, 1–19.

Lehmann, R. (2020). The forecasting power of the ifo business survey. CESifo Working Paper, 8291.

Ma, C., Rogers, J. H., & Zhou, S. (2020). Global economic and financial effects of 21st century pandemics and epidemics. SSRN. http://dx.doi.org/10.2139/ssrn.3565646

Mitze, T., Kosfeld, R., Rode, J., & Wälde, K. (2020). Face masks considerably reduce COVID-19 cases in germany: A synthetic control method approach. IZA Discussion Paper, 13319.

Prather, K. A., Wang, C. C., & Schooley, R. T. (2020). Reducing transmission of SARS-CoV-2. Science, 368, 1422–1424.

Prem, K., Liu, Y., Russell, T. W., Kucharski, A. J., Eggo, R. M., Davies, N., Flasche, S., Clifford, S., Pearson, C. A., Munday, J. D., Et al. (2020). The effect of control strategies to reduce social mixing on outcomes of the covid-19 epidemic in wuhan, china: A modelling study. The Lancet Public Health, e261–e270.

Robert Koch Institute, (2020). Corona virus disease 2019 (COVID-19) daily situation report of the Robert Koch Institute. Robert Koch Institut. Retrieved June 19, 2020, from https://www.rki.de/DE/Content/InfAZ/N/NeuartigesCoronavirus/Situationsberichte/2020-06-17-en.pdf?_blob=publicationFile

Sauer, S., & Wohlrabe, K. (2020). Ifo Handbuch der Konjunkturumfragen. ifo Beiträge zur Wirtschaftsforschung.

Vanella, P., Wiessner, C., Holz, A., Krause, G., Moehl, A., Wiegel, S., Lange, B., & Becher, H. (2020). The role of age distribution, time lag between reporting and death and healthcare system capacity on case fatality estimates of COVID-19. ResearchSquare. https://doi.org/10.21203/rs.3.rs-38592/v1

Wollmershäuser, T., Göttert, M., Grimme, C., Krolage, C., Lautenbacher, S., Lehmann, R., Link, S., Rathje, A.-C., Reif, M., Sandqvist, A. P., Et al. (2020). ifo Konjunkturprognose Sommer 2020: Deutsche Wirtschaft–es geht wieder aufwärts. ifo Schnelldienst, 73(1), 3–58.

